# Primary care and point-of-care testing during a pandemic: Clinician’s perspectives on integrating rapid testing for COVID-19 into the primary care pathway

**DOI:** 10.1101/2021.04.13.21255347

**Authors:** Patrick Kierkegaard, Timothy Hicks, Yaling Yang, Joseph Lee, Gail Hayward, Philip J. Turner, A. Joy Allen, Brian D. Nicholson

## Abstract

**Background:** Real-world evidence to support the adoption of SARS-CoV-2 point-of-care (POC) tests in primary care is limited. As the first point of contact of the health system for most patients, POC testing can potentially support general practitioners (GPs) quickly identify infectious and non-infectious individuals to rapidly inform patient triaging, clinical management, and safely restore more in-person services.

**Objectives:** To explore the potential role of SARS-CoV-2 point-of-care testing in primary care services.

**Design:** A qualitative study using an inductive thematic analysis.

**Setting:** 21 general practices located across three regions in England.

**Results:** Three major themes were identified related to POC test implementation in primary care: (1) Insights into SARS-CoV-2 POC tests; (2) System and organisational factors; and (3) Practice-level service delivery strategies. Thematic subcategories included involvement in rapid testing, knowledge and perception of the current POC testing landscape, capacity for testing, economic concerns, resource necessities, perception of personal risk and safety, responsibility for administering the test, and targeted testing strategies.

**Conclusion:** GPs knowledge of POC tests influences their degree of trust, uncertainty, and their perception of risk of POC test use. Concerns around funding, occupational exposure, and workload play a crucial role in GPs hesitation to provide POC testing services. These concerns could potentially be addressed with government funding, the use of targeted testing, and improved triaging strategies to limit testing to essential patient cohorts.

## BACKGROUND

Testing for SARS-CoV-2 is central to the global response to COVID-19. Laboratory-based reverse transcriptase or digital polymerase chain reaction (PCR) to detect SARS-CoV-2 has been criticised for results taking too long, and for requiring specialist operators at certified laboratories [1]. Point-of-care (POC) tests promise rapid results without the need for specialised facilities but the evidence for real-world SARS-CoV-2 POC tests accuracy, implementation, and adoption is limited.

General practices play an important role in the diagnosis, treatment, and control of infectious diseases [2-4]. As the first point of contact with the health system for most patients, general practice conducts over 95% of all health system activity in the UK [5, 6]. POC tests could support general practitioners (GPs) by rapidly informing patient triage, clinical management and by facilitating in-person consultations [7]. However, GPs have well documented reservations about POC tests accuracy, over-reliance on the tests, costs in the absence of reimbursement, integration into existing clinic workflows, and their overall clinical utility [8-12].

With so many concerns about POC tests use, it is critical to take account of GPs knowledge and attitudes about POC tests for SARS-CoV-2 to enable appropriate implementation strategies. We used semi-structure interviews to explore GPs views on the impact that SARS-CoV-2 POC tests might have on general practice, to assess the feasibility of introducing them into general practice, examine the potential impact on infection control in primary care, and to identify facilitators and barriers to adoption including attitudes toward SARS-CoV-2 POC tests.

## METHODS

### Study Design

This was a qualitative study, we used semi-structured interviews to capture GPs perceptions regarding the integration of SARS-COV-2 POC tests into routine primary care services during the pandemic [13]. Our methods and results are reported in accordance with the Consolidated Criteria for Reporting Qualitative Research (COREQ) [14].

### Setting

We purposefully sampled GPs from a broad geographic distribution of general practices across three regions of England [15].

### Recruitment

With support from three NIHR Local Clinical Research Networks (LCRN), GPs were invited by a standardised email outlining the purpose of the study. Eligible participants were English speaking, practicing GPs, providing care during the pandemic. GPs who worked at practices that were closed, or not providing care services throughout the pandemic, were not eligible. Reasons for nonparticipation were not elicited. A participant information sheet, visual pathway diagram triaging SARS-COV-2 testing, and consent form were sent to GPs who expressed an interest. We did not reimburse participants. The authors had no prior contact or relationships with the majority of research participants.

We obtained written, informed consent from all study participants. Interviewees were asked for permission to record interviews, and to publish excerpts from interviews. Apart from one participant, all interviewees granted the team permission to record the interview and to publish deidentified excerpts from the interview.

The project approved and registered as a service evaluation by the Newcastle Joint Research Office and recorded on the Newcastle upon Tyne Hospital Foundation Trust’s Clinical Effectiveness Register (Project no. 10222). The Newcastle University Faculty of Medical Sciences Research Ethics Committee reviewed the protocol and deemed the work exempt from ethical approval.

### Interview topic guide

We used an interview guide to prompt study participants to share their perspectives. The interview guide was informed by prior research conducted by members of the study team [16-18], informal discussions with primary and secondary care physicians, and prior studies on the role of primary care during past epidemics [19-21]. We iteratively refined it after review by a general practitioner (BDN) and two pilot interviews. A pathway diagram representing patient triage into national SARS-CoV-2 testing centres was also included to stimulate discussion, based on information extracted from NHS and NICE guidelines (Supplemental file 1).

### Data collection

Semi-structured interviews were conducted via videoconference between September and November 2020 by an experienced male qualitative researcher (PK) and three researchers (one male, two females) with training in qualitative methods (TH, YY, JA). All interviews lasted between 30-60 minutes. We documented observations about each interview (e.g., field notes) immediately after each interview. We continued to recruit and interview study participants until no new themes emerged from the interviews (saturation) [22].

### Data Management and Analysis

Data management and analysis took place from October to December 2020. All interviews were transcribed verbatim using the Otter.ai software and both interview transcripts and notes were checked, anonymised, and corrected against the audio files. Transcripts were not returned to participants for review. Anonymized interview transcripts were securely stored on an encrypted server.

During data collection, the study team met regularly to review content and themes. We used NVivo 1.3 software (QSR International) for inductive thematic analysis [13]. Four researchers coded the transcripts: a health services researcher (PK), diagnostics evaluation methodologist (JA), biomedical engineer (TH), and health economist (YY). Transcripts were read and re-read to identify recurring themes [23]. PK coded transcripts and drafted the codebook using open coding followed by closed thematic coding to allow the iterative expansion and reduction of themes and subthemes [13]. The codebook was discussed amongst the research team in weekly meetings. Newly identified themes (intercoder agreement) relating to the original transcripts were aligned where necessary. Disagreements were resolved by consensus. Interviews continued until data saturation, determined when the study team judged that no new themes were identified [24-26]. All four researchers (PK, TH, YY, JA) then re-read the results to ensure they reflected the original interview data.

## RESULTS

### Participants

We recruited twenty-two GPs (10 women and 12 men) from twenty-one general practices across five regions in the UK (Table 1). All the participants were actively involved in providing remote and in-person care services to patients during the pandemic.

**Table 1:** Demographic features of participants and characteristics of study sites.

| **Participant characteristics** | **Number** |
| --- | --- |
| **Total number of participants** | 22 |
| **Sex** |  |
| Male | 12 |
| Female | 10 |
| **Medical training** |  |
| Average time post qualification (years), median | 18 |
| Range of qualification time (years), median | 29 |
| **Study site characteristics** |  |
| **Region of practice** |  |
| Thames Valley and South Midlands | 9 |
| London | 4 |
| North East and North Cumbria | 8 |
| **Number of patients registered to practice, mean** | 14522 (3600 - 40,000) |
| **Practice setting** |  |
| Urban | 7 |
| Suburban | 1 |
| Rural | 5 |
| Mixed | 8 |

### Themes

Our analysis revealed three major themes, and several subthemes (Table 2). Quotes are anonymized to protect participant confidentiality. The following sections describe each of these themes and summarize the key findings with illustrative quotes.

**Table 2:** Overview of themes and subthemes.

| Theme | Subthemes | Description | Illustrative quote |
| --- | --- | --- | --- |
| **Awareness of SARS-CoV-2 testing** | Involvement in rapid testing | The degree in which GPs are involved providing POC testing for SARS-CoV-2. | "We don't currently have a rapid test. I'd say there isn't current provision or not through our practice or NHS." (GP 06) |
|  | Knowledge and perception of the current POC test landscape | GPs understanding of POC tests currently in-development or approved for market use. | "I don't know anything about them. I knew that they're very early on this point of care testing for antibodies but that seems to have gone away." (GP 01) |
|  | Value to care management | The perceived value of POC testing to help in the clinical management of patients. | "Patients, especially with respiratory symptoms, would benefit from a rapid testing, because then we can actually see them, or the patients who have weak symptoms who we don't know if they have got COVID or not. So, I think it will fit in. It will be immensely helpful." (GP 10) |
| **System and organisational factors related to service implementation** | Capacity for testing | The anticipated impact of implementing POC tests and fears of overwhelming primary care services. | "Because primary care doesn't have capacity to test as a general rule." (GP 05) |
|  | Economic concerns | Factors related to costs and incentives and their relationship to workload capacity. | "We've still got to carry on trying to earn our QOF points and so there's not been any leeway there at all." (GP 19) |
| **Practice-level implementation and service delivery strategies** | Risk mitigation | Viewpoints concerning the affect POC tests would have on service delivery considering the potential for occupational exposure and assurance. | "As an individual practice, I think it's sort of the anxiety to have POC [tests] and be the contact. How would you practically administer those tests and what sort of PPE would we need to see these patients?" (GP 01) |
|  | Responsibility for administering the test | Preferences in terms of which staff member should be assigned to perform the testing. | "That's not something that my surgery will be keen to provide a GP for, because you've spent all day taking a sample. And it is something that personally, I feel that an HCA or, you know, someone of a less extensive qualification would be able to be trained to deliver" (GP 14) |
|  | Targeted testing strategies | Discussions where GPs preferred to use a selection criterion strategy to determine who should be tested. | "You would prioritise that resource to the symptomatic unwell patients .... The asymptomatic testing really would only be when you've got plenty of resource to do that because, you know, suddenly your tests are going to go through the roof." (GP 09) |
|  | Testing location | The locational set-up of where and how general practices would triage and test patients. | "I think if it's just a quick point of care test, I think more practices potentially might take it up and arrange a thing where they see the patient in the car park and do a very quick test." (GP 07) |

### Theme 1: Awareness of SARS-CoV-2 testing

#### Involvement in rapid testing

GPs reported limited exposure, experience, and access to SARS-CoV-2 POC tests. All respondents said they did not have access to POC tests.

> “There was no direct testing available in the COVID Clinic because there was no testing within primary care nationwide, as far as we’re aware, and so anybody that needed a test would be directed to the gov.uk website or 119 to get their test done.” (GP 08)

GPs said they often received inquiries from patients under the impression that they could access SARS-CoV-2 tests.

> “We have several people a day asking for tests and we have several people asking why we can’t do the tests, and we give them the same answer every time that we don’t have access to the tests.” (GP 16)

#### Knowledge of current POC testing landscape

GPs general level of understanding of POC tests varied across practices. In most cases, their knowledge of SARS-CoV-2 POC tests was based on the news or social media leading several of them to express concerns around the limited evidence-base.

> “Not a great deal because we don’t really have much information. I know that there’s a swab related [test], what we would call a pregnancy test, or a lateral flow test. But that’s basically all what we know.” (GP 14)

Although GPs familiarity with these tests varied, there was a broad consensus that the evidence-base for POC tests and their accuracy needs to be improved to raise their confidence in using the tests.

> “I know that the technology is out there, I don’t know how accurate it is, and how easy it is to use. By that I mean, it doesn’t sound like very reliable sources. But that is the most, I think that’s the quickest way you can get information and you have to be very careful about how you assess whether the data is credible or not.” (GP 01)

#### Value to care management

GPs expressed that POC tests could potentially add value to the management of patient care, especially in terms of supporting them in distinguishing between COVID-19 and other respiratory illnesses to inform effective triaging, and treatment decisions.

> “I think it will definitely help because if you get a result quite quickly, at least that way you can reassure yourself and the patient quite quickly that they don’t have COVID and don’t need to self-isolate. You can tell them “You’ve got a chest infection, here are your antibiotics”. I think that’s that would help a lot.” (GP 07)

Another anticipated that POC testing would enable them to restore in-person care management for other patient cohorts, particularly those with other respiratory illnesses.

> “Patients, especially with respiratory symptoms, would benefit from a rapid testing, because then we can actually see them, or the patients who have weak symptoms who we don’t know if they have got COVID or not.” (GP 10)

Finally, some GPs said that that tests would be useful for cases of opportunistic testing in the event a patient visits the practice for another condition but exhibits COVID-19 symptoms.

> “They’ve come with a skin infection on their leg cellulitis and they come down to the practice and it’s quite clear when you’re seeing them that their temperature may well be due to cellulitis. But they’ve also got a cough. They’ve also lost their taste and smell, and they’ve just been distracted by the cellulitis. You’re sitting in front of a patient in a surgery who might have COVID. Now, it makes no sense at all, for me to send that patient away to go to a regional test site, I want to test that patient there.” (GP 21)

### Theme 2: System and organisational factors related to service implementation

#### Capacity for testing

GPs were concerned that offering POC tests would increase attendance and affect continuity of care by drawing resources away from patients with chronic disease or urgent clinical needs.

> “One of the problems is, in terms of all the other stuff, that primary care has to deal with all the other normal cancer and heart disease and stuff. If primary care gets overwhelmed with testing, it’s struggling as it is coping with demand, so it adds an extra layer of demand.” (GP 12) Most of the respondents emphasised that already busy general practices would struggle to handle the potentially large influx of patients requesting SARS-CoV-2 testing.
>
> “I don’t think it would be able to work through our system because it would overload it… and we know, we can’t just manage doing that.” (GP 15)

A few GPs considered that testing should be offered based on regional disease prevalence. They were reluctant to provide testing if local prevalence was high but prepared to if local prevalence was low.

> “If we’ve got a high prevalence, it’s not something we’ve got the capacity to deal with… But for very low prevalence, speaking like any other standard test I need to complete, I’ll be using it with my trained healthcare assistant.” (GP 15)

#### Economic concerns

The GPs interviewed indicated that POC tests could add pressure onto general practices who already need to meet targets to generate income. GPs expressed that they would need additional resources to hire extra staff in order to minimise the disruption of existing services to meet their reporting requirements.

> “I think because there’s still an expectation for practices to meet all the targets for everything … is there a way to get another healthcare assistant? For example, a nurse or a GP running this separately with which you know at least that way it wouldn’t impact on current services that are having to happen.” (GP 07)

One general practitioner mentioned that general practices would willingly adopt POC tests if financial incentives were introduced to perform the testing.

> “If you monetize the process, we will look at it…. I think if you monetize this and set up a protocol, a lot of GPs will look at it.” (GP 01)

There were also concerns about the additional costs relation to procuring infection prevention easures.

> “You’ve got extra cleaning fees for the room… glass screens at reception are thousands of pounds.” (GP 15)

### Theme 3: Practice-level implementation and service delivery strategies

#### Risk mitigation

Some GPs felt that negative results from POC tests could reassure staff and patients that they are at a reduced risk of exposure during face-to-face consultations.

> “I think it definitely would make us feel safer again and I think more importantly it would make other patients feel safe because we do still have patients who are very frightened about coming to use our facilities.” (GP 19)

The use of POC tests in general practice could also give GPs more confidence to invite the patients into the clinic, given they were presently reluctant to offer a face-to-face assessment.

> “I think it will improve (and) it will make us more confident in face-to-face consultations. So, we’ve got a huge population with respiratory illness, especially COPD. I think these are the patients who kind of have been missed out on getting seen.” (GP 10)

However, GPs also explained that there was an increased risk of staff anxiety and absence if there was increased risk of occupational exposure to potentially infectious individuals booking appointments to get tested.

> “Some people wouldn’t come into work, because they would say it’s not safe for them to come into work.” (GP 17)

#### Responsibility for administering the test

Health care assistants (HCAs) or nurse practitioners were identified by most interview participants as the most suitable and cost-effective GPs to administer POC tests.

> “It’s a skill that needs to be learned, but it’s quite a simple one. You need someone who’s focused on just that one problem. But it’s also time consuming. So, nurse practitioners but they are a lot more expensive. So, you want someone who’s not gonna be huge cost and resources.” (GP 15)

Participants reported that this would also ensure that GPs could devote their limited face-to-face time with patients to provide clinical care as opposed to performing POC tests.

> “I’d see it probably more being a nurse or a healthcare assistant if it’s just the point-of-care test. It’s all about kind of using skills appropriately, isn’t it? And obviously trying to free up the doctor time for more doctory things really.” (GP 09)

#### Targeted testing strategies

Several GPs suggested that testing resources should be reserved for use based on clinical need. They felt that testing should be allocated to unwell patients.

> “It should be at the discretion of GPs to test when they feel that it is clinically necessary for patient care… For people who don’t need any clinical input, I don’t want to see 50 patients lined up in the morning to have a COVID-19 test. This should not be an alternative to the drive thru testing or the walking testing. It should be for safe patient care, where they actually need to be seen.” (GP 10)

There was a consensus amongst participants that testing should only be reserved for patients who are considered high-risk or vulnerable, require in-person consultation, and are unable to travel to a testing centre.

> “I’d choose the high-risk groups first, but the frail people who can’t travel and a lot of anxious people with lung conditions. Probably people who’ve got mobility problems, difficulties getting to test centres.” (GP 15)

#### Testing location

GPs said that testing should be conducted outside to reduce the risk of infection inside the practice.

> “If it’s a point-of-care test, maybe something even in a car park, where you’ve got someone driving through and you do a test and they drive off with the result straightaway if possible, or you phone them back if something’s longer” (GP 07)

A few respondents said that if there was a high volume of testing, POC tests should be offered at a separate mass testing site to reduce the risk of transmission.

> “I think there should be a separate site rather than the general practice where they can get that rapid test, just to reduce potential risk of cross infection.” (GP 09)

However, some GPs believed that testing patients in a hub nearby the clinic would be the most appropriate option.

> “We’re talking about like a porta cabin or something separate somewhere from the building by you, we can see our own patients, or we open up our red hub again, and we can see our own patients, because we’re the only ones that really know them.” (GP25)

## DISCUSSION

We found multiple challenges to adoption of SARS-CoV-2 POC tests in primary care. GPs had significant reservations based on the limited information available, an overwhelming workload, and the potential for increased occupational exposure to SARS-CoV-2. GPs were more likely to adopt POC tests if conducting testing added value to care management, if additional resources were made available to offset the increased workload, and evidence was available to assure them that POC tests would reduce occupational and patient exposure.

### Comparison to the existing literature

Occupational exposure was a concern amongst the GPs we interviewed as increased interaction with patients would entail staff being exposed to more potentially infectious individuals. Increased occupational exposure to patients has been linked with increased stress and anxiety amongst healthcare workers related to being infected and infecting their families [27-32]. Without establishing and communicating the risks associated with the introduction of POC tests, GPs may not feel reassured by testing, which they fear could result in staff absenteeism during a pandemic [33-39]. Education and pandemic response training may mitigate fear and absenteeism among clinicians [40, 41].

GPs had limited awareness of the SARS-CoV-2 POC tests. Although GPs were somewhat familiar with some types of POC tests, most of their understanding was sourced from various mass media, and they report concerned with the lack of robust ‘real-world’ evidence [42-45]. This suggests that GPs current attitudes and expectations of SARS-CoV-2 POC tests are shaped by a combination of knowledge gaps, perceived risks, and uncertainties. These factors are consequential as they are critical determinants that inform decisions and behaviours [46-49].

There was a consensus amongst interview participants that HCAs should administer POC tests. Part of this motivation was that HCAs are cost-effective, and are accustomed to taking on responsibilities that remove burdens from GPs and nurses [50, 51]. This reasoning resonates with previous work exploring team-based models in primary care focused on the redistribution of tasks among care team members. Optimising workforce capacity by re-delegating tasks to non-medically qualified staff members can help GPs and nurses can prioritise unwell patients requiring treatment [52-57]. However, we did not seek the views of HCAs.

### Implications for research and practice

It may be necessary to incentivise POC tests in primary care [58-61]. GPs argued that increasing capacity to deliver POC tests is dependent on securing additional funding to create the necessary infrastructure. Alternative time-limited funding arrangements may be needed to increase capacity for POC tests use, such as the Alternative Provider Medical Services (APMS) contract, to cover all associated costs until the pandemic has passed [62]. Workforce optimisation strategies to share resources between general practices across the ‘primary care network’ (PCN) could offset workload burden for individual practices [63]. However, this may not be a sustainable approach across England given the high variability between PCNs organisational structures and characteristics [64]. GPs suggested that outdoor testing stations would facilitate infection control and reduce the need for patients to travel to distant community testing sites [65], especially frail patients, and or those without vehicles [66]. GPs expressed a strong preference for modular buildings, portable cabins or tents in practice carparks that could be used to triage and dispatch patients and assess contamination risk. This approach is supported by evidence from secondary care from both the COVID-19 and previous SARS pandemic [67-69].

GPs indicated that POC testing could add value to patient management if it served as a discriminator between SARS-CoV-2 and other respiratory viruses. For instance, reliably distinguishing between COVID-19 and influenza clinically is impossible because of the overlap of clinical presentations [70-72].

This suggests that POC testing that can facilitate syndromic testing could provide value in guiding clinical management. Thus, future developments for SARS-CoV-2 POC tests could help meet GPs clinical needs of distinguishing between respiratory illnesses by focusing on multiplex testing. Related studies in secondary care suggests that multiplex testing for influenza and RSV can a positive impact on patient management and is associated with more appropriate clinical decisions, reduced antibiotic use, timelier infection control measures, more appropriate antiviral management, and reduced costs in secondary care settings [73-77].

POC tests are less efficient and more error-prone when handled by non-laboratory trained individuals [78, 79]. For instance, the accuracy of a lateral flow immune assay for SARS-CoV-2 dropped significantly when used by non-laboratory trained professionals [80]. As GPs identified HCAs as the ideal candidates to administer the POC tests this raises questions about the training required to ensure they are

administered correctly. We suggest studies developing training protocols and standard operating procedures for individuals without medical or laboratory backgrounds to minimize inaccurate results and test failures [81]. The role of POC tests in home visiting and out-of-hour services warrants further investigation [82] [83].

### Strengths and Limitations

A strength was the qualitative methods we used. They allowed us to explore the views and experiences of general practice staff in an in-depth and descriptive manner. The topic guide was piloted with two GPs who were not participants in the study, with minimal changes recommended. Although the sample size was small, we achieved information saturation appropriate to a qualitative study design when no new themes were discovered during the interviews [22]. Concurrent thematic analysis ensured that data saturation occurred before data collection was complete.

A limitation is that we included general practices from only three regions of England, which may not have captured the variation in clinical practice and might therefore limit the generalisability of findings. The interview participants did not include any nurses and HCAs, who are likely to have play a central part of POC use in primary care. Lastly, the interviews occurred between the 25th of September 2020 and the 27th of October when the COVID-19 situation in the UK was changing rapidly, immediately prior to the second national lockdown in November 2020. It is possible participants priorities may have changed subsequently.

## CONCLUSION

We explored the perspectives of general practice staff on adopting SARS-CoV-2 POC tests into clinical routine practice. Our findings highlight that GPs awareness of the POC testing landscape varies, as do their perception of risk, and uncertainty regarding the adoption of these tests. Our interviews revealed concerns around the support general practices would need to manage the additional workload, staffing, and risks of occupational exposure. General practices would be willing to provide point-of-care testing services if these concerns could be mitigated through increased government funding, and targeted testing and triaging strategies to limit testing services to essential patient cohorts. Our findings provide important information that can inform policy development concerning planning and implementation of mass testing programmes for COVID-19 and future pandemics.

## Supporting information

COREQ Checklist

## Data Availability

All relevant data are within the manuscript and its supporting information files.

## Abbreviations

(BMA): British Medical Association
(CCG): Clinical commissioning group
(COVID-19): Coronavirus Disease 2019
(GP): General Practitioner
(HCA): Health care assistant
(LCRN): Local Clinical Research Network
(LMC): Local Medical Committees
(NIHR): National Institute of Health Research
(POC): Point of care
(QOF): Quality and Outcomes Framework
(SARS-CoV-2): Severe acute respiratory syndrome coronavirus 2

## Acknowledgements

The authors would like to thank the general practices who participated in the study. The study team is also grateful to the NHIR Clinical Research Networks for assisting in the recruitment of the study sites. The authors thank Prof. Peter Buckle for helping review the first draft of the paper and providing valuable feedback and to Dr Sara Graziadio for helpful advice on the initial draft of the interview topic guide. We are grateful to Drs Kile Green, Sam G. Urwin, Jana Suklan, Clare Lendrem and Amanda Winter for double checking the accuracy of the transcripts. We are also thankful for the advice and feedback provided by the NIHR Newcastle MIC Insight Panel.

## Author contributions

PK drafted the manuscript. AJA, TH, YY, BN, PJT, GH, JL contributed to the editing and provided critical feedback. PK, TH, YY, AJA, BN designed the study. PK, TH, YY, AJA collected and analysed the data. All authors reviewed and approved the final version of the manuscript.

## Data availability statement

All relevant data are within the manuscript and its supporting information files.

